## Supplementary File S2: Supplementary Figures S1, S2 for "*Plasmodium falciparum hrp2* and *hrp3* gene deletion status in Africa and South America by highly sensitive and specific digital PCR"

Claudia A. Vera-Arias *et al.*

### Supplementary File S2

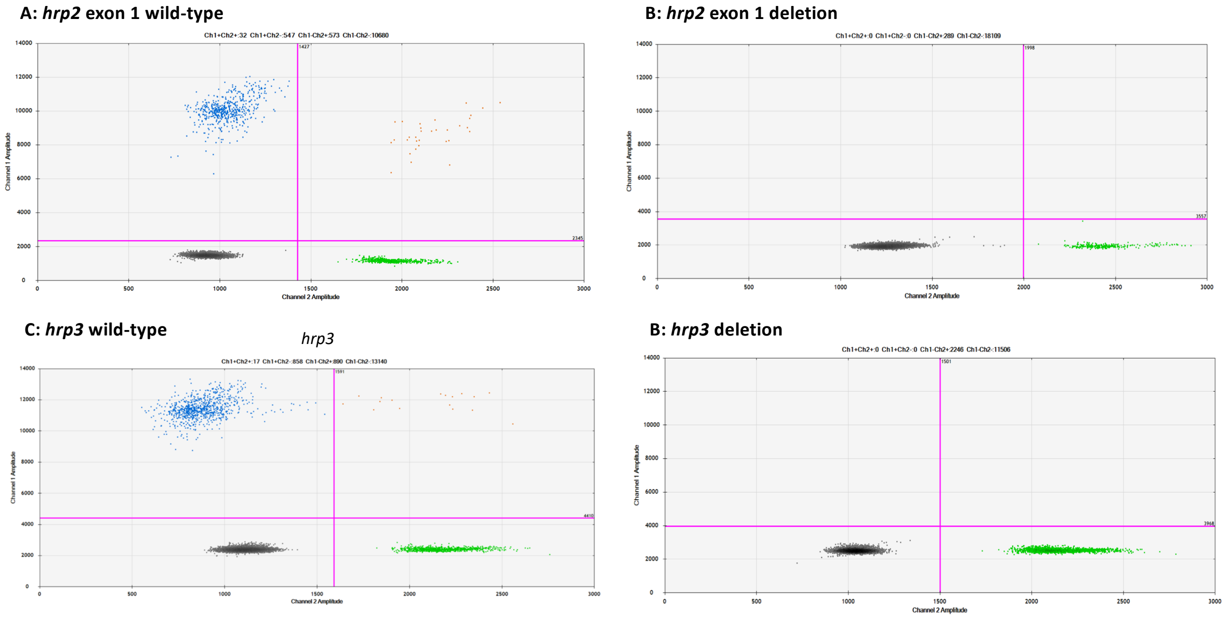


**Supplementary Figure S1**: Examples of *hrp2* exon 1 and *hrp3* ddPCR assays. Droplets positive for *hrp2* exon 1 or *hrp3* are shown in blue (top left of each panel). Droplets positive for *tRNA* are shown in green (bottom right). Droplets positive for *hrp2* exon1 or *hrp3* and *tRNA* are shown in orange (top right). Negative droplets are shown in gray (bottom left). A) *hrp2* exon 1 assay. Wild-type infection of medium density. Approximately 550 droplets are positive each for *hrp2* exon 1 and *tRNA*, and 32 for both targets. B) *hrp2* exon 1 deletion. 289 droplets are positive for *tRNA*, but no droplets are positive for *hrp2* exon 1. C) *hrp3* assay. Wild-type infection of medium density. Approximately 880 droplets are positive each for *hrp3* and *tRNA*. D) *hrp3* deletion. 2246 droplets are positive for *tRNA*, but no droplets are positive for *hrp3*.

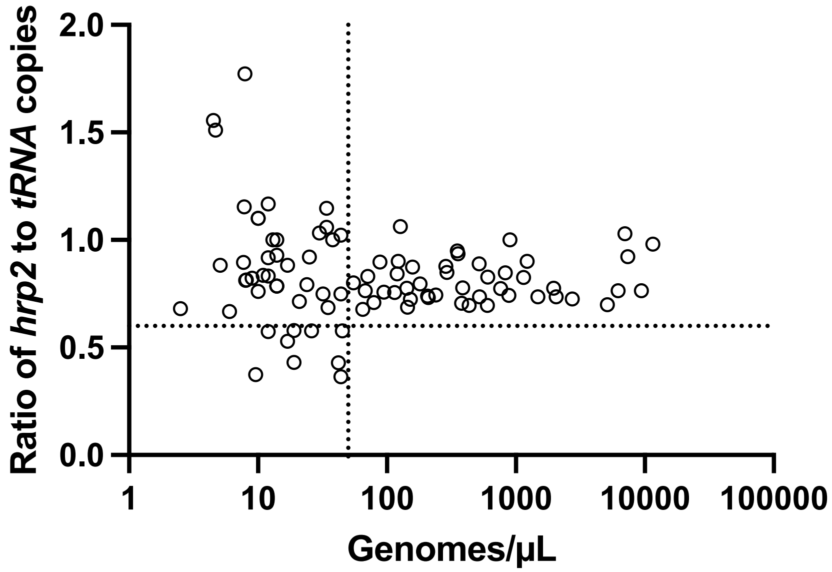


**Supplementary Figure S2:** Ratios of *hrp2* to *tRNA* copies in field isolates from Zanzibar (n=91). With increasing parasite density (X-axis), the ratio converges around 0.8. No deletions were observed in this sample set. Dashed lines show a ratio of *hrp2* to *tRNA* copies of 0.6, and 50 genomes/uL. Despite the ratio being lower than 1, mixed infection can be reliably detected at densities >50 genomes/µL, and if >40% of parasites carry the deletion.
