## Supplementary File S1: Assay conditions for "*Plasmodium falciparum hrp2* and *hrp3* gene deletion status in Africa and South America by highly sensitive and specific digital PCR"

Claudia A. Vera-Arias *et al.*

### Supplementary File S1

### ddPCR assays

Supermix from BioRad
All primers at 10 µM

***hrp2*** **exon 2 ddPCR**

| | [uL] |
|---|---|
| SuperMix for Probes (no dUTP) | 11 |
| hrp2_Exon 2_fwd | 1.76 |
| hrp2_Exon 2_rev | 1.76 |
| hrp2_Exon 2_Probe | 0.88 |
| tRNA_fwd | 0.44 |
| tRNA_rev | 0.44 |
| tRNA_Probe | 0.22 |
| H2O | 3.5 |
| DNA | 2 |

Cycling conditions

| | |
|---|---|
| 95° | 10 min |
| 94° | 30 sec |
| 56° | 1 min => 44 x back to step 2 |
| 98° | 10 min |
| 4° | hold |

***hrp2*** **exon 1 ddPCR**

| | [uL] |
|---|---|
| SuperMix for Probes (no dUTP) | 11 |
| hrp2-exon1_fwd | 1.76 |
| hrp2-exon1_rev | 1.76 |
| hrp2-exon1_Probe | 0.88 |
| tRNA_fwd | 0.44 |
| tRNA_rev | 0.44 |
| tRNA_Probe | 0.22 |
| H2O | |
| DNA | 2 |

Cycling conditions

| | |
|---|---|
| 95° | 10 min |
| 94° | 30 sec |
| 54° | 1 min => 44 x back to step 2 |
| 98° | 10 min |
| 4° | hold |

***hrp3*** **ddPCR**

| | [uL] |
|---|---|
| SuperMix for Probes (no dUTP) | 11 |
| hrp3_fwd | 0.66 |
| hrp3_rev | 0.66 |
| hrp3_Probe | 0.44 |
| tRNA_fwd | 0.44 |
| tRNA_rev | 0.44 |
| tRNA_Probe | 0.22 |
| H2O | 3.24 |
| DNA | 6 |

Cycling conditions

| | |
|---|---|
| 95° | 10 min |
| 94° | 30 sec |
| 55° | 1 min => 44 x back to step 2 |
| 4° | hold |

**Nested PCR assays**

***hrp2* nested PCR**

Polymerase, buffer, and dNTPs from Solis Biodyne
All primers at 10 µM

| **Primary PCR** | [uL] | Cycling conditions | |
|---|---|---|---|
| Taq FirePol | 0.15 | | |
| Buffer B | 1.5 | 94° | 10 min |
| MgCl2 | 1.5 | 94° | 50 sec |
| dNTPs | 1.3 | 55° | 50 sec => 39x back to step 2 |
| Primer hrp2_F1 | 0.2 | 70° | 1 min |
| Primer hrp2_R1 | 0.2 | 70° | 5 min |
| H2O | 8.25 | | |
| DNA | 2 | | |

| **Nested PCR** | **[uL]** | Cycling conditions | |
|---|---|---|---|
| Taq FirePol | 0.15 | | |
| Buffer B | 1.5 | 94° | 10 min |
| $MgCl_2$ | 1.5 | 94° | 50 sec |
| dNTPs | 1.3 | 55° | 50 sec => 39x back to step 2 |
| Primer hrp2_F2 | 0.2 | 70° | 1 min |
| Primer hrp2_R1 | 0.2 | 70° | 5 min |
| H2O | 8.25 | | |
| DNA | 2 | | |

**Primer sequences**

| | |
|---|---|
| Hrp2 F1 | 5'-CAAAAGGACTTAATTTAAATAAGAG-3' |
| Hrp2 R1 | 5'-AATAAATTTAATGGCGTAGGCA-3' |
| Hrp2 F2 | 5'-ATTATTACACGAAACTCAAGCAC-3' |

***msp2* nested PCR**

Polymerase, buffer, and dNTPs from Solis Biodyne
All primers at 10 µM

| **Primary PCR** | [uL] | Cycling conditions | |
|---|---|---|---|
| ddH2O | 7.85 | | |
| Buffer B | 1.5 | 95˚ | 1 min |
| MgCl2 25 mM | 1.5 | 95˚ | 30 sec |
| dNTPs 2.5 mM | 1.2 | 55˚ | 30 sec |
| Primer *msp2*_S2-fw | 0.4 | 72˚ | 1 min => 34 x back to step 2 |
| Primer *msp2*_S3-rev | 0.4 | 72˚ | 5 min |
| Taq FirePol | 0.15 | | |
| DNA | 2 | | |

| **Nested PCR** | [uL] | Cycling conditions | |
|---|---|---|---|
| ddH2O | 8.975 | | |
| Buffer B | 1.5 | 95˚ | 1 min |
| MgCl2 25 mM | 1.5 | 95˚ | 30 sec |
| dNTPs 2.5 mM | 1.2 | 55˚ | 30 sec |
| Primer M5_Fc27 (10 uM) | 0.15 | 72˚ | 1 min => 34 x back to step 2 |
| Primer N5 3D7 (10 uM) | 0.225 | 72˚ | 5 min |

| | |
|---|---|
| Primer S1-tail fw | 0.3 |
| Taq FirePol | 0.15 |
| Primary PCR as template | 1 |

**Primer sequences**

| | |
|---|---|
| *msp2*_S2-fw: | 5’- GAAGGTAATTAAAACATTGTC-3’ |
| *msp2*_S3-rev: | 5’- GAGGGATGTTGCTGCTCCACAG -3’ |
| *msp2*_S1Tail-fw : | 5’- GCTTATAATATGAGTATAAGGAGAA -3’ |
| *msp2_FC27 type*_M5-rev: | 5’- GCATTGCCAGAACTTGAA -3’ |
| *msp2_3D7 type*_N5 -rev: | 5’- VIC-CTGAAGAGGTACTGGTAGA -3’ |
